# Assessing the Accuracy of the NeuroBooster Array for *GBA1* Variant Detection

**DOI:** 10.64898/2026.11.10.26367159

**Authors:** Marco Toffoli, Laurel Screven, Aaron Jeffries, Raquel Real, Georgia Pittwood, Amelia Anderson, Selen Yalkic, Finlay Fraser, Jakub Stepniak, Ilaria Palmieri, Paola Dimartino, Enza Maria Valente, Global Parkinson’s Genetics Program (GP2), Lara M Lange, Cornelis Blauwendraat, Huw R Morris, Anthony HV Schapira

**Affiliations:** Department of Clinical and Movement Neurosciences, Queen Square Institute of Neurology, University College London, London, UK; Global Parkinson’s Genetics Program (GP2), Chevy Chase, MD, USA; Biosciences, Faculty of Health and Life Sciences, University of Exeter, Exeter, EX4 4QD, UK; Movement Disorders Centre, UCL Queen Square Institute of Neurology, London, UK; Area of Neurodegenerative and Adult Rare Diseases, IRCCS Mondino Foundation, Pavia, Italy; Department of Molecular Medicine, University of Pavia, Pavia, Italy; Laboratory of Neurogenetics, National Institute on Aging, Bethesda, Maryland, USA; Coalition for Aligning Science, Chevy Chase, MD, USA

## Abstract

*GBA1* variants are major Parkinson’s disease risk factors, but their detection is challenging. We compared NeuroBooster Array (NBA) genotyping with reference sequencing in 3,744 individuals. NBA correctly identified 520/608 carriers (85.5%). Of 89 missed calls, 73 had no matching probe and 16 had missing calls. 25 participants had false positive calls. NBA should not be used alone for clinical decisions, but is very useful for large case control analysis.

---

*GBA1* variants are the most important genetic risk factor for Parkinson’s disease (PD) and cause the autosomal recessive disorder Gaucher disease (GD). Accurate detection of *GBA1* risk and pathogenic variants is critical for understanding the genetics of PD, and has crucial implications for prognostication, counselling and trial eligibility^1^. However, identifying *GBA1* variants remains challenging due to the high sequence homology between exons 9-11 of *GBA1* and its pseudogene (*GBAP1*). This homology frequently leads to misalignment errors and miscalls, especially for relatively frequent severe variants such as p.L483P (also known as L444P) and RecNciI^2^. There are different approaches that can be applied for identifying variants in *GBA1*, including nested PCR of *GBA1* amplicons^3^, targeted long-read sequencing, and short-read whole-genome sequencing (WGS) with a specific bioinformatics pipeline (e.g., Gauchian)^2,4,5^. While cross-platform analyses have shown that these methodologies are effective in studying *GBA1* variation, they may be expensive and labour intensive. For large-scale projects comprising hundreds of thousands of individuals, it is highly desirable to have a high-throughput, low-cost assay for *GBA1* variant detection. To address this need, the NeuroBooster Array (NBA)^6^ was developed by the Global Parkinson’s Genetics Program (GP2)^7^ to genotype the Illumina Global Diversity backbone with a custom probe set to identify variants of interest for Parkinson’s and other neurodegenerative diseases. Here, we compare NBA *GBA1* genotyping with reference methodologies to assess NBA genotyping accuracy and missing call rate, and we provide probe metrics and the best probe for the detected *GBA1* variants.

Participants came from three cohorts: PD-Frontline (PDF), PDGEN, and the Parkinson’s Precision Medicine Initiative (PPMI). Of 3,744 samples with reference *GBA1* calls and NBA data available, all passed QC (PPMI: 2,758; PDF: 738; PDGEN: 248). Based on GP2 ancestry, 2,939 participants (78.5%) were of European and 658 (17.6%) of Ashkenazi Jewish ancestry; the remaining 147 (3.9%) belonged to nine other ancestry groups. Based on the recorded diagnosis in GP2, the 3,744 participants included 2,042 individuals with PD, 1,615 without PD (including 845 prodromal PD), and 87 classified as “Other” (Supplementary Table 1). *GBA1* variants were detected in 608 participants (16.2%): 294 (14.4%) PD cases, 306 non-PD (18.9%), and 8 “Others” (9.2%). Allele frequencies (AF) are reported in Supplementary Table 2. These AF should be interpreted considering the different recruitment strategies and enrichment for *GBA1* carriers in the three cohorts.

Across the region of interest, NBA contained 423 probes, covering 316 unique genomic positions (Supplementary Table 3).

Of the 608 carriers identified using reference sequencing, 520 (85.5%) were correctly detected using NBA genotype data. The 88 *GBA1* variant carriers who were not correctly identified with NBA (14.5% of all variant carriers and 2.4% of the whole cohort) accounted for 89 missed variant calls: 73 involved variants for which NBA had no probe targeting the variant allele (Supplementary Table 4), and 16 had a matching probe that returned no call in that individual. No carrier was missed when a matching probe returned a call.

Across all NBA probes targeting protein-altering or canonical splice-site changes in the *GBA1* region, NBA reported false positive calls in 25 participants (0.7% of all samples analysed), involving 17 variants (Supplementary Table 5). In 9 of these 25 participants, the false positive call was made concordantly by all two or three probes targeting the variant.

The best probe for each variant is reported in Table 1 and detailed metrics for each probe targeting the variants in our analysis panel are in Supplementary Table 6. Notably, the best probe for p.N409S (chr1:155235843:T:C), based on sensitivity, false positives, coverage and missingness, was not able to identify any of the 18 homozygous carriers (which were reported as heterozygotes), while another probe with poorer overall performance (ilmnseq_rs76763715.1_F2BT) was able to correctly call 17 of them. No probe identified the single p.E365K homozygote present in the dataset.

**Table 1.** Best probe per variant.

| Variant | NBA probe name | Chr1 position (hg38) | REF | ALT | TP | FP | TN | FN | missing | sensitivity* | specificity* | precision* |
|---|---|---|---|---|---|---|---|---|---|---|---|---|
| **p.R535H** | rs75822236 | 155235002 | C | T | 4 | 0 | 3736 | 0 | 4 | 100.0% | 100.0% | 100.0% |
| **c.1505+1G>T** | chr1:155204985:G:T | 155235194 | C | A | 0 | 0 | 3743 | 0 | 1 |  | 100.0% |  |
| **p.R502C** | Seq_rs80356771.1_ilmnrev_ilmnF2BT | 155235196 | G | A | 4 | 1 | 3738 | 0 | 1 | 100.0% | 100.0% | 80.0% |
| **p.V499L** | Seq_rs369068553.1_ilmnrev_ilmnF2BT | 155235205 | C | G | 1 | 0 | 3735 | 0 | 8 | 100.0% | 100.0% | 100.0% |
| **p.A495P** | 1:155205008-G-C | 155235217 | C | G | 0 | 0 | 3709 | 0 | 32 |  | 100.0% |  |
| **p.D482N** | exm106204 | 155235256 | C | T | 0 | 1 | 3743 | 0 | 0 |  | 100.0% | 0.0% |
| **c.1388+1G>A** | chr1:155205471:G:A | 155235680 | C | T | 0 | 0 | 3741 | 0 | 3 |  | 100.0% |  |
| **p.D448H** | rs1064651 | 155235727 | C | G | 0 | 0 | 801 | 0 | 4 |  | 100.0% |  |
| **p.V433L** | rs80356769 | 155235772 | C | A | 1 | 0 | 3743 | 0 | 0 | 100.0% | 100.0% | 100.0% |
| **p.E427K** | seq_rs149171124 | 155235790 | C | T | 1 | 2 | 3741 | 0 | 0 | 100.0% | 99.9% | 33.3% |
| **p.N409S** | chr1:155235843:T:C | 155235843 | T | C | 308 | 0 | 3435 | 0 | 1 | 100.0% | 100.0% | 100.0% |
| **p.T408M** | seq_rs75548401 | 155236246 | G | A | 80 | 2 | 3662 | 0 | 0 | 100.0% | 99.9% | 97.6% |
| **p.M400I** | Variant49117 | 155236269 | C | T | 0 | 2 | 3742 | 0 | 0 |  | 99.9% | 0.0% |
| **p.R368C** | Variant49120 | 155236367 | G | A | 0 | 0 | 804 | 0 | 1 |  | 100.0% |  |
| **p.E365K** | kgp308887 | 155236376 | C | T | 118 | 1 | 3623 | 0 | 2 | 100.0% | 100.0% | 99.2% |
| **p.T336S** | Neuro-1:155206254 | 155236463 | T | A | 0 | 1 | 3742 | 0 | 1 |  | 100.0% | 0.0% |
| **p.T306I** | Variant49123 | 155237423 | G | A | 1 | 0 | 3742 | 0 | 1 | 100.0% | 100.0% | 100.0% |
| **p.R296X** | chr1:155207245:C:T | 155237454 | G | A | 0 | 0 | 3743 | 0 | 1 |  | 100.0% |  |
| **p.F255Y** | 1:155207367-T-A | 155237576 | A | T | 0 | 0 | 3743 | 0 | 1 |  | 100.0% |  |
| **c.762-2A>G** | chr1:155207371-155207371_T_C | 155237580 | T | C | 0 | 0 | 3743 | 0 | 1 |  | 100.0% |  |
| **p.G241R** | Seq_rs409652 | 155238174 | C | T | 0 | 0 | 3706 | 0 | 18 |  | 100.0% |  |
| **p.G234E** | rs74462743 | 155238194 | C | T | 0 | 1 | 3742 | 0 | 1 |  | 100.0% | 0.0% |
| **p.W223R** | Variant49133 | 155238228 | A | G | 1 | 0 | 3613 | 0 | 110 | 100.0% | 100.0% | 100.0% |
| **p.P217S** | GBA_P217S | 155238246 | G | A | 0 | 1 | 3743 | 0 | 0 |  | 100.0% | 0.0% |
| **p.S212X** | chr1:155208051:C:G_ilmnrev_ilmnF2BT | 155238260 | G | C | 0 | 0 | 3743 | 0 | 1 |  | 100.0% |  |
| **p.K196Q** | rs121908297 | 155238519 | T | G | 0 | 0 | 3743 | 0 | 1 |  | 100.0% |  |
| **p.D179H** | rs147138516 | 155238570 | C | G | 1 | 0 | 3742 | 0 | 1 | 100.0% | 100.0% | 100.0% |
| **p.R170L** | rs80356763 | 155238596 | C | A | 0 | 1 | 3739 | 0 | 4 |  | 100.0% | 0.0% |
| **p.R170C** | rs398123530 | 155238597 | G | A | 0 | 0 | 3742 | 0 | 2 |  | 100.0% |  |
| **p.Q112X** | chr1:155209527:C:T | 155239736 | G | A | 0 | 0 | 3743 | 0 | 1 |  | 100.0% |  |
| **p.R87W** | 1:155209725-C-T | 155239934 | G | A | 0 | 1 | 3742 | 0 | 1 |  | 100.0% | 0.0% |
| **p.R86Q** | exm106249 | 155239936 | C | T | 1 | 0 | 3743 | 0 | 0 | 100.0% | 100.0% | 100.0% |
| **p.R83C** | Neuro-1:155209737 | 155239946 | G | A | 0 | 4 | 3740 | 0 | 0 |  | 99.9% | 0.0% |
| **p.R78C** | Neuro-1:155209752 | 155239961 | G | A | 0 | 1 | 3738 | 0 | 5 |  | 100.0% | 0.0% |
| **p.C62Y** | rs145888253 | 155240008 | C | T | 0 | 1 | 3743 | 0 | 0 |  | 100.0% | 0.0% |
| **p.G49S** | Seq_rs760930573 | 155240048 | C | T | 1 | 0 | 3743 | 0 | 0 | 100.0% | 100.0% | 100.0% |
| **p.L29Afs\*18** | IlmnSeq_rs387906315_ilmndup4 | 155240660 | D | I | 8 | 0 | 3736 | 0 | 0 | 100.0% | 100.0% | 100.0% |
| **p.K13R** | 1:155210498-TC | 155240707 | T | C | 1 | 2 | 3723 | 0 | 1 | 100.0% | 99.9% | 33.3% |

REF: reference allele. ALT: alternate allele. REF and ALT are given on the GRCh38 forward strand; *GBA1* is on the minus strand, so cDNA alleles are their complement. TP: true positives. FP: false positives. TN: true negatives. FN: false negatives. Missing: missing calls at that position. Sensitivity and precision are not given for variants with no carriers with a non-missing call.
\*Metrics are calculated only on non-missing calls.

Access to sequencing and genotyping resources is instrumental in advancing our understanding of PD, and GP2 has enabled unprecedented access to genetic information from different populations across the world. In this work, we show that NBA can correctly detect 85.5% of *GBA1* carriers, confirming its value for high-volume case-control analysis. On the other hand, a proportion of *GBA1* carriers were missed (14.5% of all *GBA1* variant carriers and 2.4% of the whole cohort). Most variants are missed because dedicated probes were not included in the current genotyping array design, but some samples also have missing calls due to underperforming probes. When a probe targeting the correct allele returned a call, no carrier was missed.

Our cohort included mainly individuals of European and Ashkenazi Jewish ancestries and had a relatively high prevalence of p.N409S carriers. The NBA probes for p.N409S showed an excellent performance overall, so it is possible that the rate of missed variants in an unselected PD cohort might be higher.

Notably, most of the variants missed in our cohort are p.L483P, the most common severe *GBA1* variant in Europeans and Asians, and the intronic variant rs3115534-G, the principal African-ancestry *GBA1* PD risk variant^8^. Missing these two variants has significant implications for studies estimating the impact of *GBA1* variants on PD pathogenesis. The potential for mislabelling participants also precludes the use of genotyping with NBA or similar for patient stratification in interventional studies, and clinicians with access to NBA data from study participants should refrain from diagnosing *GBA1* status based on NBA alone.

We provide metrics for all probes targeting the variants in our analysis panel and define the best probe for each variant. In most cases, the difference between probes is trivial, but on large datasets small differences in sensitivity and specificity might produce significant effects. False positive calls were rare, but in nine participants they were supported by all probes targeting the variant. For these variants, we cannot exclude miscalling by the reference method.

Several NBA probes share a genomic position with a clinically relevant *GBA1* variant but test a different allele, and some rsIDs (e.g. rs104886460) are multiallelic. Matching probes to variants by position or rsID alone can therefore attribute calls to the wrong variant. This highlights that probes should be matched on position and alleles, accounting for strand. In addition, for 24 custom NBA probes, the position and alleles reported in the released data correspond to a different change from that indicated by the probe name (e.g. GBA_G232E is reported at c.694, p.G232R, rather than c.695, p.G232E). The p.G232E and p.L500P carriers in our cohort were called homozygous reference by these probes (Supplementary Table 7).

Another important point is the rate of variants not called due to missingness, despite a QC step to exclude samples with missingness above 5%. This is because genome-wide call-rate QC does not protect against locus-specific genotyping failure.

The detection of p.N409S requires a specific mention. Only one probe correctly identified the majority of p.N409S homozygotes, but another probe had slightly better overall performance (when sensitivity was calculated as a presence/absence metric). Relying on one or the other probe depends on whether one wants to prioritise high sensitivity or enable the detection of p.N409S homozygotes.

In our analysis, we did not include imputed data. In certain populations, the intronic variant rs3115534-G, p.E365K and p.T408M can be imputed from NBA (as opposed to p.L483P), so imputation may improve overall detection of *GBA1* variants.

GP2 now also includes a substantial volume of WGS data, which, when properly analysed using Gauchian, can detect most or all *GBA1* variants. We encourage researchers using GP2 data to acknowledge the limitations of NBA when genotyping *GBA1* and to consider validating their findings or using WGS data whenever possible.

## Methods

All cohorts had NBA genotyping available through GP2, with each cohort using a distinct reference method to validate *GBA1* variant calls. The study methodology is shown in Figure 1.

**Figure 1:**
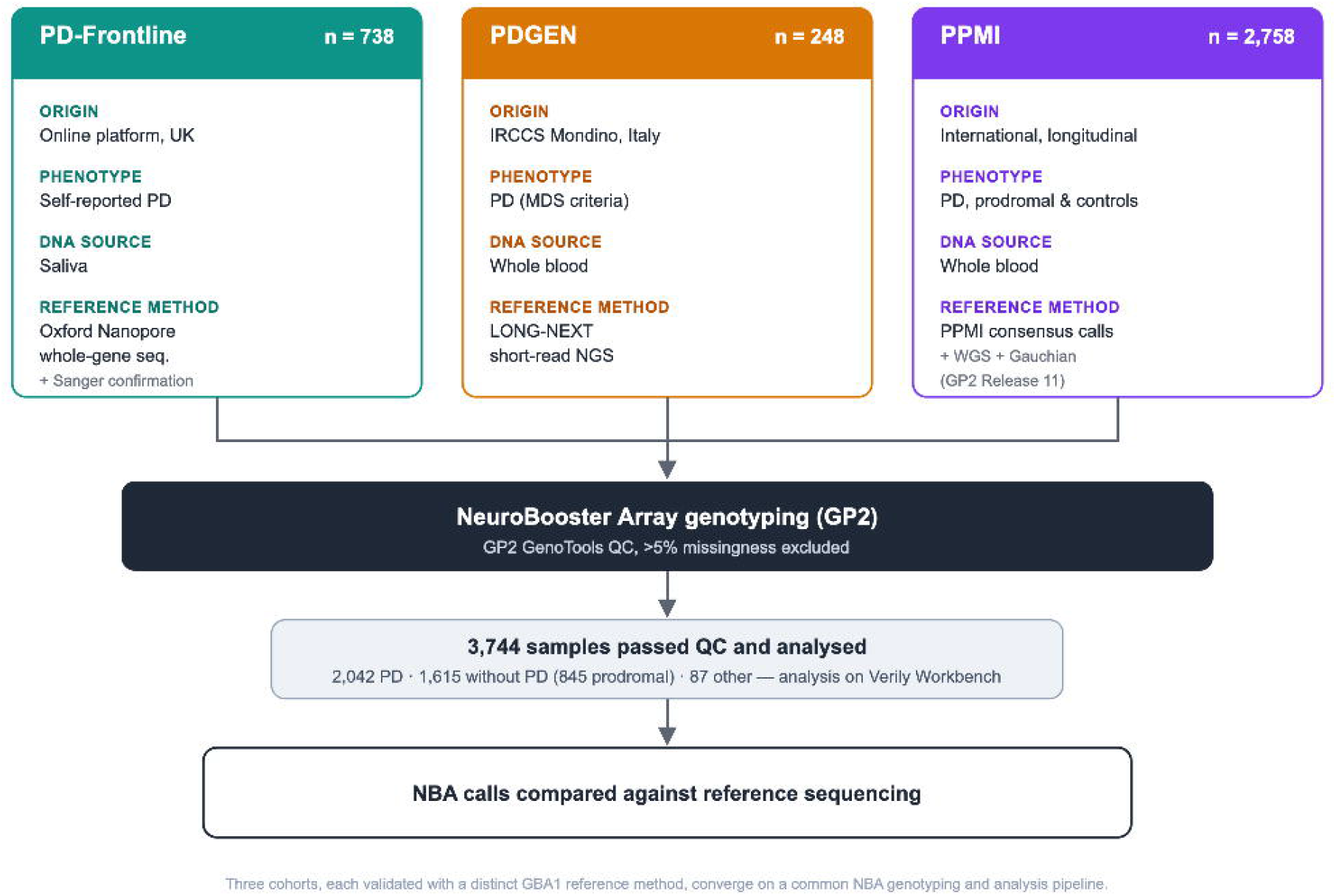
Structure of the study.

### PD-Frontline

PDF is an online platform for identifying potential candidates for clinical trials (pdfrontline.com; London, Queen Square Research Ethics Committee ref 15/LO/1155, NCT06151002). All participants had a self-reported diagnosis of PD and resided in the UK. After signing an online consent form, participants donated a saliva sample with GeneFix saliva collection kit (Oragene). DNA was used for targeted whole-gene *GBA1* sequencing with Oxford Nanopore Technologies (ON), as previously described^9^. In brief, a PCR to amplify an 8.9kb region containing the whole *GBA1* gene was followed by ON sequencing of multiplexed and barcoded samples. All *GBA1* variants identified were confirmed with Sanger sequencing of the exon of interest with nested PCR.

### PDGEN

Patients were recruited at the IRCCS Mondino Foundation (Pavia), and PD diagnosis was made by a movement disorders specialist according to the Movement Disorders Society criteria^10^. Written informed consent for genetic testing with diagnostic and research purposes was obtained from all patients. For all analyses, the input DNA was extracted from whole peripheral blood. Samples were sequenced using LONG-NEXT (LN), an NGS approach tailored for detecting *GBA1* variants via short-read sequencing^4^. This uses long-range PCR to amplify the whole *GBA1* gene in a single ∼6kb amplicon, followed by a customized bioinformatics pipeline that masks the pseudogene sequence during alignment. The PDGEN study was approved by the Regional Ethics Committee comitato etico pavia, protocollo nr. 0108640/21 del 23/12/2021.

### PPMI

PPMI (https://www.ppmi-info.org/) is an international longitudinal study that recruits individuals with manifest PD, prodromal PD, and healthy controls. PPMI includes a genetically enriched recruitment arm that enrolls individuals based on their known genetic status. Genetic consensus files downloaded from the PPMI data sharing portal on 26 August 2025 served as the reference dataset for *GBA1* variant validation. In addition, short-read WGS data generated through GP2 (Release 11; https://doi.org/10.5281/zenodo.17753486) were analysed. GP2 WGS data processing and quality control have been described previously^11^, and *GBA1* variants were called using the Gauchian algorithm, as detailed previously^8^.

### NeuroBooster Array and quality control

DNA samples obtained from the three cohorts were submitted to GP2 for genotyping using the NBA array^6^. Samples underwent standard GP2 quality control, including exclusion in case of mismatch between reported and genetic sex, as a part of the GenoTools pipeline, as described elsewhere^11^. Samples with genotype missingness above 5% across the full genotyping array content were excluded.

### Data analysis

NBA calls were obtained from GP2 Release 11 and analysis was carried out on Verily Workbench. The *GBA1* region of interest (including the 39 amino acid leader peptide) was defined as NC_000001.11:g.155234452_155244627. Only exonic variants, splicing variants and the known pathogenic intronic variant rs3115534-G were included in this analysis.

Synonymous variants were excluded and carriers of synonymous variants were considered as wild type. The analysis panel comprised all variants identified by the reference methods plus additional known pathogenic *GBA1* variants with NBA probes (Supplementary Table 6). Sensitivity, specificity and precision were calculated per probe on non-missing calls. When more than one probe was available for each variant, the best probe was defined using a four-level ranking, applied in order: highest sensitivity, fewest false positives, most coverage, fewest missing. False positives were assessed at all NBA probes in the region targeting missense, nonsense, canonical splice-site (±1–2 bp) or coding indel changes, by annotating each probe from its position and alleles (GRCh38, NM_000157.4).

### Statistics and reproducibility

Each probe was evaluated on all participants with a non-missing call: true positives (TP) and false negatives (FN) were participants carrying the variant according to the reference methods who were called non-reference or reference by the probe, respectively; false positives (FP) and true negatives (TN) were non-carriers called non-reference or reference. Sensitivity was calculated as TP/(TP+FN), specificity as TN/(TN+FP) and precision as TP/(TP+FP). Carrier detection was assessed per participant and variant. No statistical tests were performed. Analyses were performed in Python using pandas and pgenlib, and PLINK 2.0 (v2.0.0-a.6.9) was used to extract genotypes; all code is available as described in Code availability.

### Gene nomenclature

Exonic variants are referred to by their amino acid change (reference NP_000148.2) and intronic variants by their cDNA change (reference NM_000157.4, MANE Select).

### Use of AI tools

Figure 1 was created with the assistance of Claude (Anthropic).

## Data availability

Data used in this article were obtained from the Global Parkinson’s Genetics Program (GP2; https://gp2.org). We used TIER 2 data from GP2 Release 11 (https://doi.org/10.5281/zenodo.17753486). GP2 data access can be requested through AMP PD (https://amp-pd.org). PPMI data used in this article were obtained on 26th August 2025 from the PPMI database (www.ppmi-info.org/access-data-specimens/download-data), RRID: SCR_006431. For information on the study, visit www.ppmi-info.org.

Sequencing data generated with ON and LN cannot be shared publicly, as participant consent does not cover public data sharing. Participant-level *GBA1* calls from reference sequencing and NBA for all participants are provided in Supplementary Data 1.

## Code availability

All code generated for this article, and the identifiers for all software programs and packages used, are available on GitHub (https://github.com/GP2code/NBA_GBA1_accuracy) and were given a persistent identifier via Zenodo DOI: 10.5281/zenodo.22933901.

## Acknowledgements

This project was supported as part of the Global Parkinson’s Genetics Program (GP2) (https://gp2.org), a research program of Aligning Science Across Parkinson’s (ASAP) (https://ror.org/03zj4c476). ASAP is managed by the Coalition for Aligning Science (CAS) (https://ror.org/00rf4ez40). The Michael J. Fox Foundation for Parkinson’s Research (MJFF) (https://ror.org/03arq3225) is the funding and strategic partner. For a complete list of GP2 members see https://doi.org/10.5281/zenodo.7904831.

This study was also supported by Uzi and Rachel Kattan and family Fund for Parkinson’s disease research (Royal Free Charity grant 285).

The Cure Parkinson’s Trust and Parkinson’s UK support PD-Frontline.

PPMI – a public-private partnership – is funded by the Michael J. Fox Foundation for Parkinson’s Research and funding partners, including AbbVie, Alamar Biosciences, Aligning Science Across Parkinson’s (ASAP), Arrowhead Pharma, Arvinas, AskBio, BIAL, BioArctic, Biohaven, BlueRock Therapeutics, Bristol Myers Squibb, Calico Labs, Capsida Biotherapeutics, Critical Path Institute, DaCapo Brainscience, Denali, Edmond J. Safra Foundation, Eli Lilly, Gain Therapeutics, GE Healthcare, Genentech, GSK, Insitro, Johnson & Johnson Innovative Medicine, Lundbeck, Merck, Neumora, Neuron23, Novartis, Olink, Regeneron, Roche, Sanofi, Tenvie, UCB, Vanqua Bio, Voyager Therapeutics, The Weston Family Foundation.

The funders had no role in study design, data analysis and interpretation, or writing of the manuscript.

## Author contributions

MT, LS, RR, EMV, LML, CB, HRM and AHVS designed the study. MT, AJ, GP, AA, SY, FF, JS, IP, PD, LML and CB contributed to study execution. MT, IP, PD, LML and CB analysed the data. MT wrote and edited the manuscript. All authors reviewed the manuscript and read and approved the final version.

