## Supplementary Information for "Assessing the Accuracy of the NeuroBooster Array for *GBA1* Variant Detection"

**Supplementary Table 1:** Phenotype distribution across the three cohorts

| Phenotype | PDGEN | PD-Frontline | PPMI | TOTAL |
|---|---|---|---|---|
| **PD** | 212 | 29 | 864 | 1105 |
| **Affected_PD** | 16 | 645 | 276 | 937 |
| **Prodromal** | 0 | 0 | 845 | 845 |
| **Unaffected** | 2 | 63 | 414 | 479 |
| **Control** | 2 | 0 | 289 | 291 |
| **Other** | 16 | 0 | 70 | 86 |
| **Affected_Other** | 0 | 1 | 0 | 1 |
| **Total** | 248 | 738 | 2758 | 3744 |

(baseline GP2 phenotype, GP2 release-11 master key).

**Supplementary Table 2:** Allele frequencies of *GBA1* variants by cohort (according to reference sequencing)

| Variant | PDGEN | | | PD-Frontline | | | PPMI | | | Total | | | |
|---|---|---|---|---|---|---|---|---|---|---|---|---|---|
| | PD | Non-PD | Other | PD | Non-PD | Other | PD | Non-PD | Other | PD | Non-PD | Other | Grand total |
| **N (samples)** | 228 | 4 | 16 | 674 | 63 | 1 | 1140 | 1548 | 70 | 2042 | 1615 | 87 | 3744 |
| **p.N409S** | 4 (0.88%) | 2 (25.00%) | 0 (0.00%) | 12 (0.89%) | 2 (1.59%) | 1 (50.00%) | 96 (4.21%) | 207 (6.69%) | 2 (1.43 %) | 112 (2.74 %) | 211 (6.53%) | 3 (1.72 %) | 326 (4.35%) |
| **p.E365K** | 0 (0.00%) | 0 (0.00%) | 0 (0.00%) | 25 (1.85%) | 3 (2.38%) | 0 (0.00%) | 40 (1.75%) | 51 (1.65%) | 1 (0.71 %) | 65 (1.59 %) | 54 (1.67%) | 1 (0.57 %) | 120 (1.60%) |
| **p.T408M** | 4 (0.88%) | 0 (0.00%) | 1 (3.12%) | 22 (1.63%) | 1 (0.79%) | 0 (0.00%) | 27 (1.18%) | 22 (0.71%) | 3 (2.14 %) | 53 (1.30 %) | 23 (0.71%) | 4 (2.30 %) | 80 (1.07%) |
| **p.L483P** | 10 (2.19%) | 0 (0.00%) | 0 (0.00%) | 6 (0.45%) | 1 (0.79%) | 0 (0.00%) | 12 (0.53%) | 6 (0.19%) | 0 (0.00 %) | 28 (0.69 %) | 7 (0.22%) | 0 (0.00 %) | 35 (0.47%) |
| **rs3115534-G** | 0 (0.00%) | 0 (0.00%) | 0 (0.00%) | 2 (0.15%) | 0 (0.00%) | 0 (0.00%) | 3 (0.13%) | 10 (0.32%) | 0 (0.00 %) | 5 (0.12 %) | 10 (0.31%) | 0 (0.00 %) | 15 (0.20%) |
| **p.L29Afs*18** | 0 (0.00%) | 0 (0.00%) | 0 (0.00%) | 1 (0.07%) | 1 (0.79%) | 0 (0.00%) | 4 (0.18%) | 2 (0.06%) | 0 (0.00 %) | 5 (0.12 %) | 3 (0.09%) | 0 (0.00 %) | 8 (0.11%) |
| **c.115+1G>A** | 0 (0.00%) | 0 (0.00%) | 0 (0.00%) | 2 (0.15%) | 0 (0.00%) | 0 (0.00%) | 3 (0.13%) | 1 (0.03%) | 0 (0.00 %) | 5 (0.12 %) | 1 (0.03%) | 0 (0.00 %) | 6 (0.08%) |

| | | | | | | | | | | | | | |
|---|---|---|---|---|---|---|---|---|---|---|---|---|---|
| **p.R159W** | 0 (0.00%) | 0 (0.00%) | 0 (0.00%) | 0 (0.00%) | 0 (0.00%) | 0 (0.00%) | 2 (0.09%) | 2 (0.06%) | 0 (0.00%) | 2 (0.05%) | 2 (0.06%) | 0 (0.00%) | 4 (0.05%) |
| **p.R502C** | 0 (0.00%) | 0 (0.00%) | 0 (0.00%) | 3 (0.22%) | 0 (0.00%) | 0 (0.00%) | 1 (0.04%) | 0 (0.00%) | 0 (0.00%) | 4 (0.10%) | 0 (0.00%) | 0 (0.00%) | 4 (0.05%) |
| **p.R535H** | 0 (0.00%) | 0 (0.00%) | 0 (0.00%) | 0 (0.00%) | 2 (1.59%) | 0 (0.00%) | 0 (0.00%) | 2 (0.06%) | 0 (0.00%) | 0 (0.00%) | 4 (0.12%) | 0 (0.00%) | 4 (0.05%) |
| **RecNcil** | 0 (0.00%) | 0 (0.00%) | 0 (0.00%) | 1 (0.07%) | 0 (0.00%) | 1 (50.00%) | 0 (0.00%) | 1 (0.03%) | 0 (0.00%) | 1 (0.02%) | 1 (0.03%) | 1 (0.57%) | 3 (0.04%) |
| **p.G241R** | 1 (0.22%) | 0 (0.00%) | 0 (0.00%) | 1 (0.07%) | 0 (0.00%) | 0 (0.00%) | 0 (0.00%) | 0 (0.00%) | 0 (0.00%) | 2 (0.05%) | 0 (0.00%) | 0 (0.00%) | 2 (0.03%) |
| **p.R170C** | 2 (0.44%) | 0 (0.00%) | 0 (0.00%) | 0 (0.00%) | 0 (0.00%) | 0 (0.00%) | 0 (0.00%) | 0 (0.00%) | 0 (0.00%) | 2 (0.05%) | 0 (0.00%) | 0 (0.00%) | 2 (0.03%) |
| **p.W223R** | 1 (0.22%) | 0 (0.00%) | 0 (0.00%) | 0 (0.00%) | 0 (0.00%) | 0 (0.00%) | 0 (0.00%) | 1 (0.03%) | 0 (0.00%) | 1 (0.02%) | 1 (0.03%) | 0 (0.00%) | 2 (0.03%) |
| **p.G103S** | 0 (0.00%) | 0 (0.00%) | 0 (0.00%) | 1 (0.07%) | 0 (0.00%) | 0 (0.00%) | 0 (0.00%) | 0 (0.00%) | 0 (0.00%) | 1 (0.02%) | 0 (0.00%) | 0 (0.00%) | 1 (0.01%) |
| **p.G154R** | 0 (0.00%) | 0 (0.00%) | 0 (0.00%) | 0 (0.00%) | 1 (0.79%) | 0 (0.00%) | 0 (0.00%) | 0 (0.00%) | 0 (0.00%) | 0 (0.00%) | 1 (0.03%) | 0 (0.00%) | 1 (0.01%) |
| **p.E365D** | 1 (0.22%) | 0 (0.00%) | 0 (0.00%) | 0 (0.00%) | 0 (0.00%) | 0 (0.00%) | 0 (0.00%) | 0 (0.00%) | 0 (0.00%) | 1 (0.02%) | 0 (0.00%) | 0 (0.00%) | 1 (0.01%) |
| **p.D448H** | 0 (0.00%) | 0 (0.00%) | 0 (0.00%) | 1 (0.07%) | 0 (0.00%) | 0 (0.00%) | 0 (0.00%) | 0 (0.00%) | 0 (0.00%) | 1 (0.02%) | 0 (0.00%) | 0 (0.00%) | 1 (0.01%) |
| **p.D419N** | 0 (0.00%) | 0 (0.00%) | 0 (0.00%) | 1 (0.07%) | 0 (0.00%) | 0 (0.00%) | 0 (0.00%) | 0 (0.00%) | 0 (0.00%) | 1 (0.02%) | 0 (0.00%) | 0 (0.00%) | 1 (0.01%) |
| **p.A495P** | 0 (0.00%) | 0 (0.00%) | 0 (0.00%) | 0 (0.00%) | 0 (0.00%) | 0 (0.00%) | 0 (0.00%) | 1 (0.03%) | 0 (0.00%) | 0 (0.00%) | 1 (0.03%) | 0 (0.00%) | 1 (0.01%) |
| **p.D179H** | 0 (0.00%) | 0 (0.00%) | 0 (0.00%) | 0 (0.00%) | 0 (0.00%) | 0 (0.00%) | 0 (0.00%) | 1 (0.03%) | 0 (0.00%) | 0 (0.00%) | 1 (0.03%) | 0 (0.00%) | 1 (0.01%) |
| **p.F255Y** | 0 (0.00%) | 0 (0.00%) | 0 (0.00%) | 0 (0.00%) | 0 (0.00%) | 0 (0.00%) | 0 (0.00%) | 1 (0.03%) | 0 (0.00%) | 0 (0.00%) | 1 (0.03%) | 0 (0.00%) | 1 (0.01%) |
| **p.F252I** | 1 (0.22%) | 0 (0.00%) | 0 (0.00%) | 0 (0.00%) | 0 (0.00%) | 0 (0.00%) | 0 (0.00%) | 0 (0.00%) | 0 (0.00%) | 1 (0.02%) | 0 (0.00%) | 0 (0.00%) | 1 (0.01%) |
| **p.E427K** | 0 (0.00%) | 0 (0.00%) | 0 (0.00%) | 1 (0.07%) | 0 (0.00%) | 0 (0.00%) | 0 (0.00%) | 0 (0.00%) | 0 (0.00%) | 1 (0.02%) | 0 (0.00%) | 0 (0.00%) | 1 (0.01%) |
| **p.K196Q** | 0 (0.00%) | 0 (0.00%) | 0 (0.00%) | 1 (0.07%) | 0 (0.00%) | 0 (0.00%) | 0 (0.00%) | 0 (0.00%) | 0 (0.00%) | 1 (0.02%) | 0 (0.00%) | 0 (0.00%) | 1 (0.01%) |
| **p.L500P** | 1 (0.22%) | 0 (0.00%) | 0 (0.00%) | 0 (0.00%) | 0 (0.00%) | 0 (0.00%) | 0 (0.00%) | 0 (0.00%) | 0 (0.00%) | 1 (0.02%) | 0 (0.00%) | 0 (0.00%) | 1 (0.01%) |
| **p.G232E** | 0 (0.00%) | 0 (0.00%) | 0 (0.00%) | 0 (0.00%) | 0 (0.00%) | 0 (0.00%) | 1 (0.04%) | 0 (0.00%) | 0 (0.00%) | 1 (0.02%) | 0 (0.00%) | 0 (0.00%) | 1 (0.01%) |
| **p.R368C** | 0 (0.00%) | 0 (0.00%) | 0 (0.00%) | 1 (0.07%) | 0 (0.00%) | 0 (0.00%) | 0 (0.00%) | 0 (0.00%) | 0 (0.00%) | 1 (0.02%) | 0 (0.00%) | 0 (0.00%) | 1 (0.01%) |

| | | | | | | | | | | | | | |
|---|---|---|---|---|---|---|---|---|---|---|---|---|---|
| **p.R296X** | 1 (0.22%) | 0 (0.00%) | 0 (0.00%) | 0 (0.00%) | 0 (0.00%) | 0 (0.00%) | 0 (0.00%) | 0 (0.00%) | 0 (0.00%) | 1 (0.02%) | 0 (0.00%) | 0 (0.00%) | 1 (0.01%) |
| **p.M319T** | 1 (0.22%) | 0 (0.00%) | 0 (0.00%) | 0 (0.00%) | 0 (0.00%) | 0 (0.00%) | 0 (0.00%) | 0 (0.00%) | 0 (0.00%) | 1 (0.02%) | 0 (0.00%) | 0 (0.00%) | 1 (0.01%) |
| **p.N58S** | 0 (0.00%) | 0 (0.00%) | 0 (0.00%) | 1 (0.07%) | 0 (0.00%) | 0 (0.00%) | 0 (0.00%) | 0 (0.00%) | 0 (0.00%) | 1 (0.02%) | 0 (0.00%) | 0 (0.00%) | 1 (0.01%) |
| **p.K13R** | 1 (0.22%) | 0 (0.00%) | 0 (0.00%) | 0 (0.00%) | 0 (0.00%) | 0 (0.00%) | 0 (0.00%) | 0 (0.00%) | 0 (0.00%) | 1 (0.02%) | 0 (0.00%) | 0 (0.00%) | 1 (0.01%) |
| **p.G49S** | 0 (0.00%) | 0 (0.00%) | 0 (0.00%) | 1 (0.07%) | 0 (0.00%) | 0 (0.00%) | 0 (0.00%) | 0 (0.00%) | 0 (0.00%) | 1 (0.02%) | 0 (0.00%) | 0 (0.00%) | 1 (0.01%) |
| **p.R86Q** | 0 (0.00%) | 0 (0.00%) | 0 (0.00%) | 1 (0.07%) | 0 (0.00%) | 0 (0.00%) | 0 (0.00%) | 0 (0.00%) | 0 (0.00%) | 1 (0.02%) | 0 (0.00%) | 0 (0.00%) | 1 (0.01%) |
| **p.R78H** | 0 (0.00%) | 0 (0.00%) | 0 (0.00%) | 1 (0.07%) | 0 (0.00%) | 0 (0.00%) | 0 (0.00%) | 0 (0.00%) | 0 (0.00%) | 1 (0.02%) | 0 (0.00%) | 0 (0.00%) | 1 (0.01%) |
| **p.T306I** | 0 (0.00%) | 0 (0.00%) | 0 (0.00%) | 1 (0.07%) | 0 (0.00%) | 0 (0.00%) | 0 (0.00%) | 0 (0.00%) | 0 (0.00%) | 1 (0.02%) | 0 (0.00%) | 0 (0.00%) | 1 (0.01%) |
| **p.S212X** | 0 (0.00%) | 0 (0.00%) | 0 (0.00%) | 0 (0.00%) | 1 (0.79%) | 0 (0.00%) | 0 (0.00%) | 0 (0.00%) | 0 (0.00%) | 0 (0.00%) | 1 (0.03%) | 0 (0.00%) | 1 (0.01%) |
| **p.S149P** | 1 (0.22%) | 0 (0.00%) | 0 (0.00%) | 0 (0.00%) | 0 (0.00%) | 0 (0.00%) | 0 (0.00%) | 0 (0.00%) | 0 (0.00%) | 1 (0.02%) | 0 (0.00%) | 0 (0.00%) | 1 (0.01%) |
| **p.V499L** | 1 (0.22%) | 0 (0.00%) | 0 (0.00%) | 0 (0.00%) | 0 (0.00%) | 0 (0.00%) | 0 (0.00%) | 0 (0.00%) | 0 (0.00%) | 1 (0.02%) | 0 (0.00%) | 0 (0.00%) | 1 (0.01%) |
| **p.V433L** | 0 (0.00%) | 0 (0.00%) | 0 (0.00%) | 0 (0.00%) | 1 (0.79%) | 0 (0.00%) | 0 (0.00%) | 0 (0.00%) | 0 (0.00%) | 0 (0.00%) | 1 (0.03%) | 0 (0.00%) | 1 (0.01%) |
| **c.1388+1G>A** | 0 (0.00%) | 0 (0.00%) | 0 (0.00%) | 0 (0.00%) | 1 (0.79%) | 0 (0.00%) | 0 (0.00%) | 0 (0.00%) | 0 (0.00%) | 0 (0.00%) | 1 (0.03%) | 0 (0.00%) | 1 (0.01%) |
| **c.1505+1G>T** | 1 (0.22%) | 0 (0.00%) | 0 (0.00%) | 0 (0.00%) | 0 (0.00%) | 0 (0.00%) | 0 (0.00%) | 0 (0.00%) | 0 (0.00%) | 1 (0.02%) | 0 (0.00%) | 0 (0.00%) | 1 (0.01%) |
| **c.762-2A>G** | 0 (0.00%) | 0 (0.00%) | 0 (0.00%) | 0 (0.00%) | 0 (0.00%) | 0 (0.00%) | 1 (0.04%) | 0 (0.00%) | 0 (0.00%) | 1 (0.02%) | 0 (0.00%) | 0 (0.00%) | 1 (0.01%) |
| **Total (all variants)** | 31 (6.80%) | 2 (25.00%) | 1 (3.12%) | 86 (6.38%) | 14 (11.11%) | 2 (100.00%) | 190 (8.33%) | 308 (9.95%) | 6 (4.29%) | 307 (7.52%) | 324 (10.03%) | 9 (5.17%) | 640 (8.55%) |

Data are shown as allele count (frequency). Groups are obtained from GP2 Diagnosis at baseline: PD = PD + Affected_PD; Non-PD = Prodromal + Unaffected + Control; Other = Other + Affected_Other. These allele frequencies should be interpreted considering the different recruitment strategies and enrichment for *GBA1* carriers in the three cohorts. Non-PD includes controls, unaffected individuals and individuals with prodromal PD (n = 845).

**Supplementary Table 3:** List of all probes across region of interest

| CHROM | POS | REF | ALT | NBA probe name |
|---|---|---|---|---|

| | | | | |
|---|---|---|---|---|
| **1** | 155234997 | A | G | chr1:155204788:T:C |
| **1** | 155234999 | A | G | GBA_Q536R |
| **1** | 155235002 | G | A | rs75822236 |
| **1** | 155235003 | G | A | Seq_rs747506979 |
| **1** | 155235017 | G | A | GBA_T530I |
| **1** | 155235020 | A | G | chr1:155204811:A:G |
| **1** | 155235023 | A | G | chr1:155204814-155204814_T_C |
| **1** | 155235024 | A | G | Seq_rs536425950_ilmnfwd_ilmnF2BT |
| **1** | 155235027 | A | G | GBA_S527P |
| **1** | 155235057 | G | A | rs121908301 |
| **1** | 155235193 | A | T | chr1:155204984:T:A_ilmnfwd_ilmnF2BT |
| **1** | 155235193 | A | T | chr1:155204984:T:A_ilmnrev_ilmnF2BT |
| **1** | 155235193 | A | C | chr1:155204984:T:G |
| **1** | 155235194 | C | A | chr1:155204985:G:T |
| **1** | 155235195 | G | A | rs80356772 |
| **1** | 155235195 | C | G | chr1:155204986-155204986_G_C_ilmnrev_ilmnF2BT |
| **1** | 155235195 | G | A | Variant49105 |
| **1** | 155235195 | G | A | Seq_rs80356772 |
| **1** | 155235195 | C | G | chr1:155204986-155204986_G_C_ilmnfwd_ilmnF2BT |
| **1** | 155235196 | G | A | Seq_rs80356771.1_ilmnrev_ilmnF2BT |
| **1** | 155235196 | G | A | 1:155204987-C-T |
| **1** | 155235196 | G | A | rs80356771 |
| **1** | 155235196 | C | A | Seq_rs80356771.2_ilmnrev_ilmnF2BT |
| **1** | 155235202 | G | A | GBA_L500P |
| **1** | 155235205 | G | A | Seq_rs369068553.2_ilmnrev_ilmnF2BT |
| **1** | 155235205 | C | G | Seq_rs369068553.1_ilmnrev_ilmnF2BT |
| **1** | 155235205 | G | A | Variant49109 |
| **1** | 155235213 | A | T | chr1:155205004-155205004_T_A_ilmnfwd_ilmnF2BT |
| **1** | 155235213 | A | T | chr1:155205004-155205004_T_A_ilmnrev_ilmnF2BT |
| **1** | 155235217 | G | C | 1:155205008-G-C |

| | | | | |
|---|---|---|---|---|
| **1** | 155235226 | G | A | Seq_rs779958429_ilmnfwd_ilmnF2BT |
| **1** | 155235228 | G | A | Seq_rs1235609262 |
| **1** | 155235231 | A | G | 1:155205022 |
| **1** | 155235235 | A | C | chr1:155205026:A:C |
| **1** | 155235241 | G | A | Seq_rs878853317 |
| **1** | 155235243 | T | A | chr1:155205034:T:A_ilmnfwd_ilmnF2BT |
| **1** | 155235243 | T | A | chr1:155205034:T:A_ilmnrev_ilmnF2BT |
| **1** | 155235256 | G | A | seq_rs75671029 |
| **1** | 155235256 | G | A | exm106204 |
| **1** | 155235260 | G | C | Seq_rs1057519356_ilmnfwd_ilmnF2BT |
| **1** | 155235282 | A | C | chr1:155205073:T:G |
| **1** | 155235301 | G | A | Seq_rs1015281456 |
| **1** | 155235303 | A | C | Seq_rs878853320 |
| **1** | 155235307 | A | C | chr1:155205098:T:G |
| **1** | 155235311 | C | A | chr1:155205102:C:A |
| **1** | 155235312 | G | A | chr1:155205103:G:A |
| **1** | 155235312 | C | A | chr1:155205103-155205103_G_T |
| **1** | 155235314 | C | G | chr1:155205105:C:G_ilmnrev_ilmnF2BT |
| **1** | 155235314 | C | G | chr1:155205105:C:G_ilmnfwd_ilmnF2BT |
| **1** | 155235587 | G | A | chr1:155235587:C:T |
| **1** | 155235587 | G | A | Neuro-1:155205378 |
| **1** | 155235678 | C | G | chr1:155205469-155205469_C_G_ilmnfwd_ilmnF2BT |
| **1** | 155235678 | C | G | chr1:155205469-155205469_C_G_ilmnrev_ilmnF2BT |
| **1** | 155235680 | G | A | chr1:155205471:G:A |
| **1** | 155235690 | G | A | 1:155205481 |
| **1** | 155235692 | T | A | chr1:155205483:T:A_ilmnfwd_ilmnF2BT |
| **1** | 155235699 | A | G | rs74752878 |
| **1** | 155235704 | C | A | GBA_F456V |
| **1** | 155235708 | G | C | 1:155205499-C-G |
| **1** | 155235708 | G | C | Seq_rs121908295_ilmnrev_ilmnF2BT |

| | | | | |
|---|---|---|---|---|
| **1** | 155235712 | G | A | chr1:155205503:C:T |
| **1** | 155235712 | G | A | GBA_Q453R |
| **1** | 155235716 | C | A | GBA_K452Q |
| **1** | 155235721 | A | T | chr1:155205512:T:A_ilmnrev_ilmnF2BT |
| **1** | 155235721 | A | T | chr1:155205512:T:A_ilmnfwd_ilmnF2BT |
| **1** | 155235726 | T | A | 1:155205517-A-T |
| **1** | 155235726 | T | A | rs77369218 |
| **1** | 155235726 | T | A | Seq_rs77369218_ilmnrev_ilmnF2BT |
| **1** | 155235727 | C | G | ilmnseq_rs1064651 |
| **1** | 155235727 | C | G | 1:155205518-G-C |
| **1** | 155235727 | C | G | rs1064651 |
| **1** | 155235727 | C | G | ilmnseq_rs1064651_ilmndup2_ilmnbot |
| **1** | 155235744 | A | G | Seq_rs1312499220 |
| **1** | 155235747 | A | G | rs75564605 |
| **1** | 155235750 | G | A | 1:155205541-C-T |
| **1** | 155235756 | A | G | chr1:155205547-155205547_A_G |
| **1** | 155235757 | G | A | chr1:155205548:G:A |
| **1** | 155235757 | C | G | chr1:155205548:G:C_ilmnfwd_ilmnF2BT |
| **1** | 155235757 | C | A | chr1:155205548:G:T |
| **1** | 155235760 | C | A | 1:155205551-G-T |
| **1** | 155235765 | A | C | VG01S1180 |
| **1** | 155235768 | C | G | chr1:155205559:G:C_ilmnfwd_ilmnF2BT |
| **1** | 155235769 | G | A | GBA_R434C |
| **1** | 155235772 | C | A | 1:155205563-G-T |
| **1** | 155235772 | C | A | rs80356769 |
| **1** | 155235772 | C | A | ilmnseq_rs80356769 |
| **1** | 155235772 | C | A | Seq_rs80356769_ilmnfwd_ilmnF2BT |
| **1** | 155235773 | G | A | Seq_rs1484043383 |
| **1** | 155235774 | C | A | GBA_W432L |
| **1** | 155235775 | T | A | chr1:155205566:T:A_ilmnfwd_ilmnF2BT |

| | | | | |
|---|---|---|---|---|
| **1** | 155235777 | A | G | Neuro-1:155205568 |
| **1** | 155235780 | G | A | VG01S1178 |
| **1** | 155235784 | G | A | GBA_G429R |
| **1** | 155235786 | G | A | GBA_G428E |
| **1** | 155235790 | G | A | exm106213 |
| **1** | 155235790 | C | A | chr1:155205581:G:T |
| **1** | 155235790 | G | A | seq_rs149171124 |
| **1** | 155235792 | G | A | GBA_P426L |
| **1** | 155235794 | C | A | Seq_rs1026993108.2_ilmnfwd_ilmnF2BT |
| **1** | 155235794 | G | A | Seq_rs1026993108.1_ilmnfwd_ilmnF2BT |
| **1** | 155235798 | A | G | GBA_L424P |
| **1** | 155235801 | C | A | GBA_A423D |
| **1** | 155235804 | A | C | GBA_L422R |
| **1** | 155235806 | C | A | GBA_N421K |
| **1** | 155235809 | C | G | chr1:155205600:G:C_ilmnrev_ilmnF2BT |
| **1** | 155235809 | C | G | chr1:155205600:G:C_ilmnfwd_ilmnF2BT |
| **1** | 155235810 | G | A | chr1:155205601:G:A |
| **1** | 155235813 | A | C | rs77284004 |
| **1** | 155235814 | C | G | GBA_D419H |
| **1** | 155235818 | C | A | rs76014919 |
| **1** | 155235819 | G | A | Seq_rs754743440 |
| **1** | 155235820 | A | C | GBA_W417G |
| **1** | 155235823 | G | A | 1:155205614-G-A |
| **1** | 155235823 | G | A | Seq_rs121908311 |
| **1** | 155235823 | G | A | rs121908311 |
| **1** | 155235829 | C | A | Seq_rs398123528.1_ilmnfwd_ilmnF2BT |
| **1** | 155235829 | C | A | seq-rs398123528 |
| **1** | 155235829 | G | C | Seq_rs398123528.2_ilmnfwd_ilmnF2BT |
| **1** | 155235831 | A | C | chr1:155205622:A:C |
| **1** | 155235841 | C | A | chr1:155205632-155205632_G_T |

| | | | | |
|---|---|---|---|---|
| **1** | 155235841 | G | C | 1:155205632-C-G |
| **1** | 155235841 | G | C | Seq_rs121908314_ilmnfwd_ilmnF2BT |
| **1** | 155235842 | C | A | GBA_N409K |
| **1** | 155235843 | A | C | Seq_rs76763715.2_ilmnfwd_ilmnF2BT |
| **1** | 155235843 | A | C | ilmnseq_rs76763715.2_F2BT |
| **1** | 155235843 | A | G | ilmnseq_rs76763715.1_F2BT |
| **1** | 155235843 | A | G | exm106217 |
| **1** | 155235843 | A | G | chr1:155235843:T:C |
| **1** | 155236244 | G | A | chr1:155206035:G:A |
| **1** | 155236245 | G | A | rs138498426 |
| **1** | 155236246 | G | A | exm106220 |
| **1** | 155236246 | G | A | rs75548401 |
| **1** | 155236246 | G | A | seq_rs75548401 |
| **1** | 155236249 | A | G | Seq_rs1057519358 |
| **1** | 155236254 | G | C | Seq_rs75528494.2_ilmnfwd_ilmnF2BT |
| **1** | 155236254 | G | A | Seq_rs75528494.1_ilmnfwd_ilmnF2BT |
| **1** | 155236254 | C | A | Seq_rs75528494.3_ilmnfwd_ilmnF2BT |
| **1** | 155236255 | G | A | GBA_S405N |
| **1** | 155236261 | G | C | 1:155206052-G-C |
| **1** | 155236261 | G | C | Seq_rs121908307_ilmnfwd_ilmnF2BT |
| **1** | 155236262 | A | C | GBA_S403R |
| **1** | 155236264 | A | G | rs76228122 |
| **1** | 155236265 | A | G | chr1:155206056:T:C |
| **1** | 155236269 | G | A | Variant49117 |
| **1** | 155236277 | G | A | rs121908309 |
| **1** | 155236277 | G | A | Seq_rs121908309 |
| **1** | 155236282 | G | C | GBA_W396C |
| **1** | 155236285 | G | A | Seq_rs760307559 |
| **1** | 155236288 | G | A | GBA_G394D |
| **1** | 155236296 | G | A | GBA_R392W |

| | | | | |
|---|---|---|---|---|
| **1** | 155236304 | G | A | chr1:155206095:C:T |
| **1** | 155236308 | G | A | GBA_E388K |
| **1** | 155236310 | A | C | Seq_rs765182863 |
| **1** | 155236310 | A | C | Variant49118 |
| **1** | 155236313 | A | G | 1:155206104 |
| **1** | 155236322 | G | A | Neuro-1:155206113 |
| **1** | 155236327 | G | A | GBA_C381Y |
| **1** | 155236328 | A | C | 1:155206119-T-G |
| **1** | 155236330 | G | A | chr1:155206121:C:T |
| **1** | 155236331 | G | A | Seq_rs781306264 |
| **1** | 155236331 | G | A | Variant49119 |
| **1** | 155236336 | G | A | chr1:155206127-155206127_G_A |
| **1** | 155236342 | A | G | chr1:155206133:T:C |
| **1** | 155236346 | G | A | GBA_L375P |
| **1** | 155236359 | C | G | 1:155206150 |
| **1** | 155236361 | A | G | GBA_F370S |
| **1** | 155236367 | G | A | Seq_rs374306700 |
| **1** | 155236367 | G | A | Variant49120 |
| **1** | 155236376 | G | A | chr1:155236376:C:T |
| **1** | 155236376 | G | A | kgp308887 |
| **1** | 155236381 | A | G | GBA_L363P |
| **1** | 155236384 | G | A | 1:155206175-C-T |
| **1** | 155236384 | G | A | rs76539814 |
| **1** | 155236384 | G | A | Seq_rs76539814 |
| **1** | 155236399 | C | A | rs78188205 |
| **1** | 155236406 | A | T | chr1:155206197-155206197_T_A_ilmnrev_ilmnF2BT |
| **1** | 155236406 | A | T | chr1:155206197-155206197_T_A_ilmnfwd_ilmnF2BT |
| **1** | 155236407 | G | C | chr1:155206198-155206198_C_G_ilmnrev_ilmnF2BT |
| **1** | 155236407 | G | C | chr1:155206198-155206198_C_G_ilmnfwd_ilmnF2BT |
| **1** | 155236409 | C | G | Seq_rs398123526.1_ilmnrev_ilmnF2BT |

| | | | | |
|---|---|---|---|---|
| **1** | 155236409 | C | G | seq-rs398123526 |
| **1** | 155236409 | G | A | Seq_rs398123526.2_ilmnrev_ilmnF2BT |
| **1** | 155236412 | G | C | chr1:155206203-155206203_G_C_ilmnfwd_ilmnF2BT |
| **1** | 155236412 | G | C | chr1:155206203-155206203_G_C_ilmnrev_ilmnF2BT |
| **1** | 155236416 | C | A | 1:155206207-G-T |
| **1** | 155236417 | G | C | chr1:155206208:G:C_ilmnfwd_ilmnF2BT |
| **1** | 155236419 | A | G | GBA_W351R |
| **1** | 155236421 | C | A | chr1:155206212:C:A |
| **1** | 155236426 | G | A | 1:155206217-C-T |
| **1** | 155236429 | A | C | chr1:155206220:T:G |
| **1** | 155236440 | A | C | chr1:155206231:T:G |
| **1** | 155236441 | A | G | rs77321207 |
| **1** | 155236444 | T | A | rs77714449 |
| **1** | 155236463 | A | T | Neuro-1:155206254 |
| **1** | 155236467 | A | G | GBA_L335L |
| **1** | 155236469 | G | A | 1:155206260 |
| **1** | 155236470 | G | A | chr1:155206261-155206261_C_T |
| **1** | 155236470 | G | C | Seq_rs754148803_ilmnfwd_ilmnF2BT |
| **1** | 155236550 | A | C | imm_1_153472965 |
| **1** | 155237340 | C | A | chr1:155207131:G:T |
| **1** | 155237341 | G | A | chr1:155207132-155207132_G_A |
| **1** | 155237343 | G | A | Seq_rs1299171778 |
| **1** | 155237357 | G | A | rs121908298 |
| **1** | 155237360 | A | G | Seq_rs1404627009 |
| **1** | 155237369 | G | A | rs79696831 |
| **1** | 155237371 | A | G | GBA_R324C |
| **1** | 155237394 | G | A | Seq_rs1264734195 |
| **1** | 155237411 | G | A | rs74731340 |
| **1** | 155237412 | A | G | GBA_S310G |
| **1** | 155237423 | G | A | Seq_rs199628072 |

| | | | | |
|---|---|---|---|---|
| 1 | 155237423 | G | A | Variant49123 |
| 1 | 155237426 | G | A | GBA_P305L |
| 1 | 155237427 | C | G | Seq_rs770796008_ilmnfwd_ilmnF2BT |
| 1 | 155237429 | G | A | rs80116658 |
| 1 | 155237430 | G | C | chr1:155207221:G:C_ilmnrev_ilmnF2BT |
| 1 | 155237433 | C | A | Seq_rs1296507371 |
| 1 | 155237438 | G | A | rs140955685 |
| 1 | 155237439 | G | C | chr1:155207230:C:G_ilmnrev_ilmnF2BT |
| 1 | 155237439 | G | A | rs374117599 |
| 1 | 155237439 | G | C | chr1:155207230:C:G_ilmnfwd_ilmnF2BT |
| 1 | 155237444 | A | G | seq-rs794727908 |
| 1 | 155237444 | A | G | Seq_rs794727908 |
| 1 | 155237446 | C | A | GBA_F298L |
| 1 | 155237453 | G | A | 1:155207244-G-A |
| 1 | 155237453 | G | A | rs78973108 |
| 1 | 155237453 | G | A | Seq_rs78973108 |
| 1 | 155237454 | G | A | chr1:155207245:C:T |
| 1 | 155237456 | A | G | chr1:155207247-155207247_T_C |
| 1 | 155237457 | G | A | chr1:155207248-155207248_G_A |
| 1 | 155237458 | A | C | Seq_rs367968666 |
| 1 | 155237458 | A | C | rs367968666 |
| 1 | 155237458 | A | C | 1:155207249-T-G |
| 1 | 155237470 | C | A | rs121908313 |
| 1 | 155237474 | C | G | Seq_rs878853321.2_ilmnfwd_ilmnF2BT |
| 1 | 155237486 | A | G | chr1:155207277:T:C |
| 1 | 155237490 | C | A | chr1:155207281:C:A |
| 1 | 155237491 | C | A | Seq_rs765583147 |
| 1 | 155237505 | G | C | Seq_rs1057519020_ilmnrev_ilmnF2BT |
| 1 | 155237513 | G | A | Seq_rs755512507 |
| 1 | 155237519 | A | T | chr1:155207310:A:T_ilmnrev_ilmnF2BT |

| | | | | |
|---|---|---|---|---|
| 1 | 155237519 | A | T | chr1:155207310:A:T_ilmnfwd_ilmnF2BT |
| 1 | 155237520 | G | A | 1:155207311 |
| 1 | 155237524 | A | C | GBA_E272D |
| 1 | 155237526 | C | A | chr1:155207317:G:T |
| 1 | 155237531 | C | G | rs76725886 |
| 1 | 155237542 | C | G | chr1:155207333:C:G_ilmnfwd_ilmnF2BT |
| 1 | 155237542 | C | G | chr1:155207333:C:G_ilmnrev_ilmnF2BT |
| 1 | 155237564 | A | G | GBA_Y259C |
| 1 | 155237570 | T | A | chr1:155207361:A:T_ilmnfwd_ilmnF2BT |
| 1 | 155237576 | T | A | 1:155207367-T-A |
| 1 | 155237576 | T | A | Seq_rs74500255_ilmnrev_ilmnF2BT |
| 1 | 155237577 | A | C | 1:155207368-T-G |
| 1 | 155237579 | G | C | Seq_rs1237637353_ilmnrev_ilmnF2BT |
| 1 | 155237580 | A | G | chr1:155207371-155207371_T_C |
| 1 | 155238133 | G | A | chr1:155207924:G:A |
| 1 | 155238139 | T | A | rs79945741 |
| 1 | 155238140 | A | C | chr1:155207931:T:G |
| 1 | 155238144 | A | G | rs121908300 |
| 1 | 155238144 | A | G | Seq_rs121908300 |
| 1 | 155238144 | A | G | 1:155207935-T-C |
| 1 | 155238145 | A | G | chr1:155207936:A:G |
| 1 | 155238150 | G | A | GBA_A249V |
| 1 | 155238154 | G | A | GBA_W248R |
| 1 | 155238164 | A | G | rs76026102 |
| 1 | 155238173 | G | A | rs77451368 |
| 1 | 155238174 | G | A | Seq_rs409652 |
| 1 | 155238176 | C | A | chr1:155207967:C:A |
| 1 | 155238185 | A | C | GBA_K237T |
| 1 | 155238186 | A | G | Seq_rs773409311 |
| 1 | 155238186 | A | G | Variant49128 |

| | | | | |
|---|---|---|---|---|
| 1 | 155238189 | G | A | GBA_L236F |
| 1 | 155238189 | G | A | GBA_L236P |
| 1 | 155238194 | G | A | rs74462743 |
| 1 | 155238196 | C | A | GBA_G234W |
| 1 | 155238201 | G | A | GBA_G232E |
| 1 | 155238202 | A | C | GBA_G232W |
| 1 | 155238206 | A | C | Variant49130 |
| 1 | 155238209 | G | A | Seq_rs75636769_ilmnrev_ilmnF2BT |
| 1 | 155238211 | A | G | GBA_A229T |
| 1 | 155238212 | C | A | rs78911246 |
| 1 | 155238214 | A | C | Variant49131 |
| 1 | 155238214 | A | C | Seq_rs381418 |
| 1 | 155238215 | A | G | rs364897 |
| 1 | 155238215 | A | G | Seq_rs364897 |
| 1 | 155238225 | G | A | GBA_L224F |
| 1 | 155238228 | A | G | Variant49133 |
| 1 | 155238233 | G | A | VG01S3114 |
| 1 | 155238234 | C | A | Seq_rs866075757.2_ilmnfwd_ilmnF2BT |
| 1 | 155238234 | G | A | Seq_rs866075757.1_ilmnfwd_ilmnF2BT |
| 1 | 155238234 | C | A | 1:155208025-GT |
| 1 | 155238240 | A | C | GBA_T219P |
| 1 | 155238242 | G | A | Seq_rs867929413 |
| 1 | 155238242 | G | A | 1:155208033-CT |
| 1 | 155238246 | G | A | GBA_P217S |
| 1 | 155238251 | C | A | GBA_A215D |
| 1 | 155238257 | A | G | GBA_L213P |
| 1 | 155238258 | G | A | GBA_L213F |
| 1 | 155238260 | C | G | chr1:155208051:C:G_ilmnrev_ilmnF2BT |
| 1 | 155238260 | C | G | chr1:155208051:C:G_ilmnfwd_ilmnF2BT |
| 1 | 155238273 | G | A | chr1:155208064:C:T |

| | | | | |
|---|---|---|---|---|
| **1** | 155238282 | G | A | Seq_rs1338037944 |
| **1** | 155238291 | G | A | Seq_rs1009850780 |
| **1** | 155238293 | A | C | rs76500263 |
| **1** | 155238302 | G | A | rs80222298 |
| **1** | 155238307 | G | A | chr1:155208098:G:A |
| **1** | 155238308 | A | G | chr1:155208099-155208099_T_C |
| **1** | 155238318 | G | C | chr1:155208109:C:G_ilmnfwd_ilmnF2BT |
| **1** | 155238516 | C | A | chr1:155208307:G:T |
| **1** | 155238518 | A | G | chr1:155208309:A:G |
| **1** | 155238519 | A | C | rs121908297 |
| **1** | 155238525 | T | A | Seq_rs886042192_ilmnrev_ilmnF2BT |
| **1** | 155238543 | G | A | chr1:155208334:C:T |
| **1** | 155238558 | A | C | chr1:155208349-155208349_A_C |
| **1** | 155238570 | C | G | rs147138516 |
| **1** | 155238579 | G | A | chr1:155208370:G:A |
| **1** | 155238581 | C | A | GBA_A175E |
| **1** | 155238583 | A | C | chr1:155208374:T:G |
| **1** | 155238587 | G | A | GBA_T173I |
| **1** | 155238588 | A | C | GBA_T173P |
| **1** | 155238589 | C | A | chr1:155208380:C:A |
| **1** | 155238596 | C | A | rs80356763 |
| **1** | 155238597 | G | A | rs398123530 |
| **1** | 155238597 | C | A | chr1:155208388-155208388_G_T |
| **1** | 155238608 | T | A | rs79796061 |
| **1** | 155238613 | G | C | Seq_rs746019841_ilmnrev_ilmnF2BT |
| **1** | 155238614 | G | A | chr1:155208405-155208405_C_T |
| **1** | 155238618 | I | D | rs398123529 |
| **1** | 155238622 | G | A | GBA_M162V |
| **1** | 155238623 | G | A | VG01S3074 |
| **1** | 155238624 | G | A | 1:155208415-C-T |

| | | | | |
|---|---|---|---|---|
| **1** | 155238626 | A | G | chr1:155208417-155208417_T_C |
| **1** | 155238629 | G | A | Seq_rs79653797.2_ilmnrev_ilmnF2BT |
| **1** | 155238629 | G | A | 1:155208420-G-A |
| **1** | 155238629 | G | C | Seq_rs79653797.1_ilmnrev_ilmnF2BT |
| **1** | 155238629 | G | A | rs79653797 |
| **1** | 155238633 | A | C | chr1:155208424-155208424_T_G |
| **1** | 155238642 | A | G | Seq_rs1280343264 |
| **1** | 155238652 | A | G | chr1:155208443:A:G |
| **1** | 155238833 | A | G | rs188328778 |
| **1** | 155239622 | G | A | GBA_E150K |
| **1** | 155239626 | C | A | chr1:155209417:C:A |
| **1** | 155239629 | C | A | GBA_F148V |
| **1** | 155239631 | A | G | GBA_Y147C |
| **1** | 155239633 | G | A | Variant49136 |
| **1** | 155239633 | G | A | Seq_rs758447515.1_ilmnfwd_ilmnF2BT |
| **1** | 155239639 | A | C | seq-rs794727708 |
| **1** | 155239639 | A | C | Seq_rs794727708_ilmnfwd_ilmnF2BT |
| **1** | 155239666 | A | G | chr1:155209457:T:C |
| **1** | 155239676 | T | A | chr1:155209467-155209467_T_A_ilmnrev_ilmnF2BT |
| **1** | 155239676 | T | A | chr1:155209467-155209467_T_A_ilmnfwd_ilmnF2BT |
| **1** | 155239678 | A | G | chr1:155209469:A:G |
| **1** | 155239685 | G | A | GBA_A129T |
| **1** | 155239691 | G | C | chr1:155209482-155209482_C_G_ilmnrev_ilmnF2BT |
| **1** | 155239699 | A | G | chr1:155209490:T:C |
| **1** | 155239705 | G | A | Seq_rs868211463 |
| **1** | 155239705 | G | A | 1:155209496-CT |
| **1** | 155239709 | C | A | 1:155209500-CA |
| **1** | 155239715 | G | A | Neuro-1:155209506 |
| **1** | 155239716 | G | C | 1:155209507-G-C |
| **1** | 155239716 | G | C | Seq_rs121908312_ilmnrev_ilmnF2BT |

| | | | | |
|---|---|---|---|---|
| **1** | 155239716 | G | C | rs121908312 |
| **1** | 155239720 | A | G | GBA_V117A |
| **1** | 155239733 | T | A | chr1:155209524:A:T_ilmnrev_ilmnF2BT |
| **1** | 155239733 | T | A | chr1:155209524:A:T_ilmnfwd_ilmnF2BT |
| **1** | 155239736 | G | A | chr1:155209527:C:T |
| **1** | 155239739 | G | A | chr1:155209530-155209530_C_T |
| **1** | 155239740 | I | D | Seq_rs779619231 |
| **1** | 155239741 | G | C | chr1:155209532-155209532_G_C_ilmnfwd_ilmnF2BT |
| **1** | 155239756 | A | G | GBA_L105P |
| **1** | 155239762 | C | A | chr1:155209553-155209553_C_A |
| **1** | 155239894 | G | A | Seq_rs757848523 |
| **1** | 155239933 | G | A | Neuro-1:155209724 |
| **1** | 155239934 | G | A | 1:155209725-C-T |
| **1** | 155239936 | G | A | exm106249 |
| **1** | 155239937 | G | A | chr1:155209728:C:T |
| **1** | 155239939 | G | A | 1:155209730-G-A |
| **1** | 155239946 | G | A | Neuro-1:155209737 |
| **1** | 155239948 | G | A | Seq_rs1141811 |
| **1** | 155239948 | G | A | Variant49138 |
| **1** | 155239951 | G | A | GBA_S81N |
| **1** | 155239957 | A | G | Neuro-1:155209748 |
| **1** | 155239961 | G | A | Neuro-1:155209752 |
| **1** | 155239967 | A | C | GBA_F76V |
| **1** | 155239971 | A | G | Seq_rs761621516_ilmnrev_F2BTindel |
| **1** | 155239972 | C | G | rs371592589 |
| **1** | 155239974 | A | G | GBA_G74S |
| **1** | 155239989 | G | A | Neuro-1:155209780 |
| **1** | 155240007 | A | C | chr1:155209798:T:G |
| **1** | 155240007 | A | G | GBA_D63N |
| **1** | 155240008 | G | A | rs145888253 |

| | | | | |
|---|---|---|---|---|
| 1 | 155240011 | A | G | chr1:155209802:A:G |
| 1 | 155240027 | C | A | chr1:155209818-155209818_G_T |
| 1 | 155240033 | C | A | 1:155209824-G-T |
| 1 | 155240038 | G | A | Seq_rs1275724188 |
| 1 | 155240048 | G | A | Variant49141 |
| 1 | 155240048 | G | A | Seq_rs760930573 |
| 1 | 155240057 | A | G | Variant49142 |
| 1 | 155240057 | A | G | Seq_rs142761046 |
| 1 | 155240064 | G | C | chr1:155209855:C:G_ilmnrev_ilmnF2BT |
| 1 | 155240064 | G | C | chr1:155209855:C:G_ilmnfwd_ilmnF2BT |
| 1 | 155240629 | A | C | rs104886460 |
| 1 | 155240630 | G | C | chr1:155210421:G:C_ilmnfwd_ilmnF2BT |
| 1 | 155240630 | G | C | chr1:155210421:G:C_ilmnrev_ilmnF2BT |
| 1 | 155240637 | G | A | Seq_rs777383151 |
| 1 | 155240649 | G | C | Seq_rs1244961782_ilmnrev_ilmnF2BT |
| 1 | 155240651 | D | I | rs80356760 |
| 1 | 155240660 | D | I | Variant2131 |
| 1 | 155240660 | D | I | IlmnSeq_rs387906315_ilmndup6 |
| 1 | 155240660 | D | I | IlmnSeq_rs387906315_ilmndup4 |
| 1 | 155240660 | D | I | IlmnSeq_rs387906315_ilmndup5 |
| 1 | 155240662 | A | C | Seq_rs387906315_ilmnfwd_F2BTindel |
| 1 | 155240672 | I | D | rs397518433 |
| 1 | 155240687 | A | G | exm1717216 |
| 1 | 155240687 | A | G | seq_rs143187997 |
| 1 | 155240707 | A | G | 1:155210498-TC |
| 1 | 155240707 | A | G | seq_rs150466109 |
| 1 | 155240711 | G | A | Seq_rs763770350 |
| 1 | 155241085 | G | A | 1:155210876-G-A |
| 1 | 155241085 | G | A | rs80356759 |
| 1 | 155241090 | C | G | Seq_rs759983265_ilmnrev_ilmnF2BT |

| | | | | |
|---|---|---|---|---|
| **1** | 155241262 | A | G | Seq_rs1232943445 |
| **1** | 155241315 | A | G | kgp33964320 |
| **1** | 155243333 | T | A | imm_1_153479748 |

REF and ALT are shown as reported in the GP2 Release 11 genotype files and are not necessarily on the GRCh38 forward strand (for forward-strand alleles of probes assigned to analysed variants, see Supplementary Table 6). D/I: deletion/insertion alleles of indel probes.

### **Supplementary Table 4:** Variants missed as NBA has no probe for them.

| Variant name | N missed due to no probe |
|---|---|
| **p.L483P** | 35 |
| **rs3115534-G*** | 14 |
| **c.115+1G>A** | 6 |
| **p.R159W** | 4 |
| **RecNciI** | 3 |
| **p.D419N** | 1 |
| **p.G103S** | 1 |
| **p.G232E** | 1 |
| **p.G154R** | 1 |
| **p.R78H** | 1 |
| **p.N58S** | 1 |
| **p.S149P** | 1 |
| **p.M319T** | 1 |
| **p.E365D** | 1 |
| **p.F252I** | 1 |
| **p.L500P** | 1 |

*This variant can be imputed using NBA data

### **Supplementary Table 5:** False positive calls by NBA

| Variant name | NBA probe(s) | N false positive calls |
|---|---|---|
| **p.R83C** | **Neuro-1:155209737** | 4 |

| | | |
|---|---|---|
| **p.E427X** | **chr1:155205581:G:T** | 3 |
| **p.E427K** | **exm106213, seq_rs149171124** | 2 |
| **p.K13R** | **1:155210498-TC, seq_rs150466109** | 2 |
| **p.M400I** | **Variant49117** | 2 |
| **p.R502C** | **1:155204987-C-T, Seq_rs80356771.1_ilmnrev_ilmnF2BT, rs80356771** | 2 |
| **p.T408M** | **exm106220, rs75548401, seq_rs75548401** | 2 |
| **p.C62Y** | **rs145888253** | 1 |
| **p.D482N** | **exm106204, seq_rs75671029** | 1 |
| **p.E365K** | **chr1:155236376:C:T, kgp308887** | 1 |
| **p.G234E** | **rs74462743** | 1 |
| **p.P217S** | **GBA_P217S** | 1 |
| **p.R170L** | **rs80356763** | 1 |
| **p.R78C** | **Neuro-1:155209752** | 1 |
| **p.R87W** | **1:155209725-C-T** | 1 |
| **p.T336S** | **Neuro-1:155206254** | 1 |
| **p.W396S** | **GBA_W396C** | 1 |

N: number of participants with a false positive call at one or more probes. All NBA probes targeting missense, nonsense, canonical splice-site or coding indel changes were assessed. p.E427X and p.W396S are called by probes not assigned to a variant present in the reference data. One p.E427X call occurred in the p.E427K carrier; the probe interrogates the same base. In 9 participants, the false positive call was made concordantly by all probes targeting the variant (two or three probes): p.R502C (n = 1), p.T408M (n = 2), p.E427K (n = 2), p.E365K (n = 1), p.D482N (n = 1) and p.K13R (n = 2).

**Supplementary Table 6:** Detailed metrics for all probes

| Variant | NBA Probe name | Chromosomal position (hg38) | REF | ALT | TP | FP | TN | FN | missing | n_truth_pos | sensitivity* | specificity* | precision* |
|---|---|---|---|---|---|---|---|---|---|---|---|---|---|
| **p.N409S** | chr1:155235843:T:C | 155235843 | T | C | 308 | 0 | 3435 | 0 | 1 | 308 | 100.0% | 100.0% | 100.0% |
| **p.N409S** | ilmnseq_rs76763715.1_F2BT | 155235843 | T | C | 278 | 0 | 3432 | 0 | 34 | 278 | 100.0% | 100.0% | 100.0% |
| **p.E365K** | kgp308887 | 155236376 | C | T | 118 | 1 | 3623 | 0 | 2 | 118 | 100.0% | 100.0% | 99.2% |
| **p.E365K** | chr1:155236376:C:T | 155236376 | C | T | 117 | 1 | 3622 | 0 | 4 | 117 | 100.0% | 100.0% | 99.2% |
| **p.T408M** | rs75548401 | 155236246 | G | A | 80 | 2 | 3656 | 0 | 6 | 80 | 100.0% | 99.9% | 97.6% |

| | | | | | | | | | | | | | |
|---|---|---|---|---|---|---|---|---|---|---|---|---|---|
| **p.T408M** | seq_rs75548401 | 155236246 | G | A | 80 | 2 | 3662 | 0 | 0 | 80 | 100.0% | 99.9% | 97.6% |
| **p.T408M** | exm106220 | 155236246 | G | A | 79 | 2 | 3662 | 0 | 1 | 79 | 100.0% | 99.9% | 97.5% |
| **p.N409S** | exm106217 | 155235843 | T | C | 77 | 0 | 3009 | 0 | 0 | 77 | 100.0% | 100.0% | 100.0% |
| **p.L29Afs*18** | IlmnSeq_rs387906315_ilmndup4 | 155240660 | D | I | 8 | 0 | 3736 | 0 | 0 | 8 | 100.0% | 100.0% | 100.0% |
| **p.L29Afs*18** | IlmnSeq_rs387906315_ilmndup5 | 155240660 | D | I | 8 | 0 | 3736 | 0 | 0 | 8 | 100.0% | 100.0% | 100.0% |
| **p.L29Afs*18** | IlmnSeq_rs387906315_ilmndup6 | 155240660 | D | I | 8 | 0 | 3735 | 0 | 1 | 8 | 100.0% | 100.0% | 100.0% |
| **p.L29Afs*18** | Seq_rs387906315_ilmnfwd_F2BTindel | 155240662 | A | C | 8 | 0 | 3734 | 0 | 2 | 8 | 100.0% | 100.0% | 100.0% |
| **p.L29Afs*18** | Variant2131 | 155240660 | D | I | 8 | 0 | 3736 | 0 | 0 | 8 | 100.0% | 100.0% | 100.0% |
| **p.R502C** | 1:155204987-C-T | 155235196 | G | A | 4 | 2 | 3738 | 0 | 0 | 4 | 100.0% | 99.9% | 66.7% |
| **p.R502C** | Seq_rs80356771.1_ilmnrev_ilmnF2BT | 155235196 | G | A | 4 | 1 | 3738 | 0 | 1 | 4 | 100.0% | 100.0% | 80.0% |
| **p.R502C** | rs80356771 | 155235196 | G | A | 4 | 1 | 3736 | 0 | 3 | 4 | 100.0% | 100.0% | 80.0% |
| **p.R535H** | rs75822236 | 155235002 | C | T | 4 | 0 | 3736 | 0 | 4 | 4 | 100.0% | 100.0% | 100.0% |
| **p.D179H** | rs147138516 | 155238570 | C | G | 1 | 0 | 3742 | 0 | 1 | 1 | 100.0% | 100.0% | 100.0% |
| **p.E427K** | exm106213 | 155235790 | C | T | 1 | 2 | 3740 | 0 | 1 | 1 | 100.0% | 99.9% | 33.3% |
| **p.E427K** | seq_rs149171124 | 155235790 | C | T | 1 | 2 | 3741 | 0 | 0 | 1 | 100.0% | 99.9% | 33.3% |
| **p.G49S** | Seq_rs760930573 | 155240048 | C | T | 1 | 0 | 3743 | 0 | 0 | 1 | 100.0% | 100.0% | 100.0% |
| **p.G49S** | Variant49141 | 155240048 | C | T | 1 | 0 | 3743 | 0 | 0 | 1 | 100.0% | 100.0% | 100.0% |
| **p.K13R** | 1:155210498-TC | 155240707 | T | C | 1 | 2 | 3723 | 0 | 1 | 1 | 100.0% | 99.9% | 33.3% |
| **p.K13R** | seq_rs150466109 | 155240707 | T | C | 1 | 2 | 3723 | 0 | 1 | 1 | 100.0% | 99.9% | 33.3% |
| **p.R86Q** | exm106249 | 155239936 | C | T | 1 | 0 | 3743 | 0 | 0 | 1 | 100.0% | 100.0% | 100.0% |
| **p.T306I** | Seq_rs199628072 | 155237423 | G | A | 1 | 0 | 3740 | 0 | 3 | 1 | 100.0% | 100.0% | 100.0% |
| **p.T306I** | Variant49123 | 155237423 | G | A | 1 | 0 | 3742 | 0 | 1 | 1 | 100.0% | 100.0% | 100.0% |
| **p.V433L** | 1:155205563-G-T | 155235772 | C | A | 1 | 0 | 3741 | 0 | 2 | 1 | 100.0% | 100.0% | 100.0% |
| **p.V433L** | Seq_rs80356769_ilmnfwd_ilmnF2BT | 155235772 | C | A | 1 | 0 | 3742 | 0 | 1 | 1 | 100.0% | 100.0% | 100.0% |
| **p.V433L** | ilmnseq_rs80356769 | 155235772 | C | A | 1 | 0 | 3739 | 0 | 4 | 1 | 100.0% | 100.0% | 100.0% |
| **p.V433L** | rs80356769 | 155235772 | C | A | 1 | 0 | 3743 | 0 | 0 | 1 | 100.0% | 100.0% | 100.0% |
| **p.V499L** | Seq_rs369068553.1_ilmnrev_ilmnF2BT | 155235205 | C | G | 1 | 0 | 3735 | 0 | 8 | 1 | 100.0% | 100.0% | 100.0% |
| **p.W223R** | Variant49133 | 155238228 | A | G | 1 | 0 | 3613 | 0 | 110 | 1 | 100.0% | 100.0% | 100.0% |

| | | | | | | | | | | | | | |
|---|---|---|---|---|---|---|---|---|---|---|---|---|---|
| **p.A495P** | 1:155205008-G-C | 155235217 | C | G | 0 | 0 | 3709 | 0 | 32 | 0 | | 100.0% | |
| **p.C62Y** | rs145888253 | 155240008 | C | T | 0 | 1 | 3743 | 0 | 0 | 0 | | 100.0% | 0.0% |
| **p.D448H** | 1:155205518-G-C | 155235727 | C | G | 0 | 0 | 87 | 0 | 0 | 0 | | 100.0% | |
| **p.D448H** | ilmnseq_rs1064651 | 155235727 | C | G | 0 | 0 | 799 | 0 | 6 | 0 | | 100.0% | |
| **p.D448H** | ilmnseq_rs1064651_ilmndup2_ilmnbot | 155235727 | C | G | 0 | 0 | 50 | 0 | 0 | 0 | | 100.0% | |
| **p.D448H** | rs1064651 | 155235727 | C | G | 0 | 0 | 801 | 0 | 4 | 0 | | 100.0% | |
| **p.D482N** | exm106204 | 155235256 | C | T | 0 | 1 | 3743 | 0 | 0 | 0 | | 100.0% | 0.0% |
| **p.D482N** | seq_rs75671029 | 155235256 | C | T | 0 | 1 | 3743 | 0 | 0 | 0 | | 100.0% | 0.0% |
| **p.F255Y** | 1:155207367-T-A | 155237576 | A | T | 0 | 0 | 3743 | 0 | 1 | 0 | | 100.0% | |
| **p.F255Y** | Seq_rs74500255_ilmnrev_ilmnF2BT | 155237576 | A | T | 0 | 0 | 3743 | 0 | 1 | 0 | | 100.0% | |
| **p.G234E** | rs74462743 | 155238194 | C | T | 0 | 1 | 3742 | 0 | 1 | 0 | | 100.0% | 0.0% |
| **p.G241R** | Seq_rs409652 | 155238174 | C | T | 0 | 0 | 3706 | 0 | 18 | 0 | | 100.0% | |
| **p.K196Q** | rs121908297 | 155238519 | T | G | 0 | 0 | 3743 | 0 | 1 | 0 | | 100.0% | |
| **p.M400I** | Variant49117 | 155236269 | C | T | 0 | 2 | 3742 | 0 | 0 | 0 | | 99.9% | 0.0% |
| **p.P217S** | GBA_P217S | 155238246 | G | A | 0 | 1 | 3743 | 0 | 0 | 0 | | 100.0% | 0.0% |
| **p.Q112X** | chr1:155209527:C:T | 155239736 | G | A | 0 | 0 | 3743 | 0 | 1 | 0 | | 100.0% | |
| **p.R170C** | rs398123530 | 155238597 | G | A | 0 | 0 | 3742 | 0 | 2 | 0 | | 100.0% | |
| **p.R170L** | rs80356763 | 155238596 | C | A | 0 | 1 | 3739 | 0 | 4 | 0 | | 100.0% | 0.0% |
| **p.R296X** | chr1:155207245:C:T | 155237454 | G | A | 0 | 0 | 3743 | 0 | 1 | 0 | | 100.0% | |
| **p.R368C** | Seq_rs374306700 | 155236367 | G | A | 0 | 0 | 803 | 0 | 2 | 0 | | 100.0% | |
| **p.R368C** | Variant49120 | 155236367 | G | A | 0 | 0 | 804 | 0 | 1 | 0 | | 100.0% | |
| **p.R78C** | Neuro-1:155209752 | 155239961 | G | A | 0 | 1 | 3738 | 0 | 5 | 0 | | 100.0% | 0.0% |
| **p.R83C** | Neuro-1:155209737 | 155239946 | G | A | 0 | 4 | 3740 | 0 | 0 | 0 | | 99.9% | 0.0% |
| **p.R87W** | 1:155209725-C-T | 155239934 | G | A | 0 | 1 | 3742 | 0 | 1 | 0 | | 100.0% | 0.0% |
| **p.S212X** | chr1:155208051:C:G_ilmnfwd_ilmnF2BT | 155238260 | G | C | 0 | 0 | 3740 | 0 | 4 | 0 | | 100.0% | |
| **p.S212X** | chr1:155208051:C:G_ilmnrev_ilmnF2BT | 155238260 | G | C | 0 | 0 | 3743 | 0 | 1 | 0 | | 100.0% | |
| **p.T336S** | Neuro-1:155206254 | 155236463 | T | A | 0 | 1 | 3742 | 0 | 1 | 0 | | 100.0% | 0.0% |
| **c.1388+1G>A** | chr1:155205471:G:A | 155235680 | C | T | 0 | 0 | 3741 | 0 | 3 | 0 | | 100.0% | |

| | | | | | | | | | | | | |
|---|---|---|---|---|---|---|---|---|---|---|---|---|
| **c.1505+1G>T** | chr1:155204985:G:T | 155235194 | C | A | 0 | 0 | 3743 | 0 | 1 | 0 | | 100.0% |
| **c.762-2A>G** | chr1:155207371-155207371_T_C | 155237580 | T | C | 0 | 0 | 3743 | 0 | 1 | 0 | | 100.0% |

REF: reference allele. ALT: alternate allele. REF and ALT are given on the GRCh38 forward strand; *GBA1* is on the minus strand, so cDNA alleles are their complement. TP: true positives. FP: false positives. TN: true negatives. FN: false negatives. Missing: missing calls at that position. n_truth_pos: number of carriers identified by the reference methods with a non-missing call at that probe (TP + FN). Sensitivity and precision are not given for variants with no carriers with a non-missing call.
*Metrics are calculated only on non-missing calls.

**Supplementary Table 7:** Custom NBA probes whose reported position and alleles correspond to a different change from that indicated by the probe name

| NBA probe name | Chromosomal position (hg38) | REF | ALT | Change at reported position | Consequence at reported position |
|---|---|---|---|---|---|
| **GBA_L500P** | 155235202 | G | A | p.L500L | synonymous |
| **GBA_F456V** | 155235704 | C | A | p.M455I | missense |
| **GBA_Q453R** | 155235712 | G | A | p.Q453X | stop gained |
| **GBA_K452Q** | 155235716 | C | A | p.Y451X | stop gained |
| **GBA_W396C** | 155236282 | G | C | p.W396S | missense |
| **GBA_R392W** | 155236296 | G | A | p.V391V | synonymous |
| **GBA_E388K** | 155236308 | G | A | p.W387X | stop gained |
| **GBA_L375P** | 155236346 | G | A | p.L375F | missense |
| **GBA_F370S** | 155236361 | A | G | p.F370L | missense |
| **GBA_W351R** | 155236419 | A | G | p.H350H | synonymous |
| **GBA_L335L** | 155236467 | A | G | p.V334V | synonymous |
| **GBA_R324C** | 155237371 | A | G | p.Q323Q | synonymous |
| **GBA_A249V** | 155238150 | G | A | p.A249T | missense |
| **GBA_W248R** | 155238154 | G | A | p.T247T | synonymous |
| **GBA_L236P** | 155238189 | G | A | p.L236F | missense |
| **GBA_G234W** | 155238196 | C | A | p.K233N | missense |

| | | | | | |
|---|---|---|---|---|---|
| **GBA_G232E** | 155238201 | G | A | p.G232R | missense |
| **GBA_G232W** | 155238202 | A | C | p.N231K | missense |
| **GBA_A229T** | 155238211 | A | G | p.G228G | synonymous |
| **GBA_M162V** | 155238622 | G | A | p.P161P | synonymous |
| **GBA_F148V** | 155239629 | C | A | p.Y147X | stop gained |
| **GBA_Y147C** | 155239631 | A | G | p.Y147H | missense |
| **GBA_G74S** | 155239974 | A | G | p.L73L | synonymous |
| **GBA_D63N** | 155240007 | A | G | p.C62C | synonymous |

REF and ALT are shown as reported in the GP2 Release 11 genotype files and are not necessarily on the GRCh38 forward strand. Change at reported position: amino acid change corresponding to the position and alleles reported in the released data. X: stop codon.

**Supplementary Data 1:** Participant-level *GBA1* calls from reference sequencing and NBA (best probe per variant) for all 3,744 participants (provided as a separate file).
Each row corresponds to one participant, identified by their GP2 ID. PDF_sequencing, PDGEN_sequencing and PPMI_sequencing report the GBA1 genotype obtained with the reference method. NBA_call reports the NBA genotype using the best probe for each variant (Table 1). 0: no variant included in the analysis was detected (synonymous variants were not reported, and carriers of synonymous variants are considered wild type).
